# Cardiovascular health in early pregnancy and circulating placental biomarkers

**DOI:** 10.1101/2024.08.16.24312132

**Authors:** Andrea C. Kozai, Bethany Barone Gibbs, Samuel Parry, Sadiya S. Khan, William Grobman, Lisa D. Levine, Rebecca McNeil, David M. Haas, Jessica L. Pippen, Robert M. Silver, Judith H. Chung, Janet M. Catov

## Abstract

**Background:** Maternal cardiovascular and placental health are related to pregnancy outcomes, yet how cardiovascular health (CVH) in early pregnancy affects placentation is not well characterized. Our objective was to estimate associations between CVH and concentrations of placental proteins. We hypothesized that more favorable CVH metrics would be associated with more favorable circulating concentrations of pregnancy-associated plasma protein A (PAPP-A), vascular endothelial growth factor (VEGF), placental growth factor (PlGF), soluble FMS-like tyrosine kinase 1 (sFlt-1), and soluble endoglin (sEng) in the 1^st^ and 2^nd^ trimesters.

**Methods:** Participants from the nuMoM2b prospective cohort with valid biomarker measures were included (n=2188). Maternal CVH was defined using the American Heart Association’s Life’s Essential 8 composite score, averaging six components measured in the 1^st^ trimester: body mass index (BMI), blood pressure, physical activity, diet, sleep duration, and smoking. Individual component scores were also evaluated. Linear regression associated scores with 1^st^ and 2^nd^ trimester circulating placental biomarkers.

**Results and Conclusions:** Higher (more favorable) composite CVH scores were associated with higher PAPP-A (favorable) in both trimesters. In contrast, higher CVH scores were associated with lower VEGF, higher sFlt-1, and higher sEng in the 1^st^ trimester (adverse). Healthier BMI scores demonstrated similar associations as those found with composite CVH. Greater physical activity was associated with more favorable 1^st^ trimester profiles (higher VEGF and lower sFlt-1), but lower 2^nd^ trimester PAPP-A. Composite CVH, potentially driven by BMI and physical activity components, may be an important factor relating to circulating concentrations of placental biomarkers in the first half of pregnancy.

## Introduction

Pregnancy is a cardiovascular stress test.^1–3^ Many adverse pregnancy outcomes (APOs) are associated with poor placental implantation and development^4–7^, but the mechanisms connecting maternal cardiovascular health (CVH), poor placental development, and APOs are still being elucidated.^8^ Risk factors for APOs are similar to those for cardiovascular disease, indicating shared pathophysiology may exist.^2^ Evidence is needed to identify those at increased risk who may benefit from early intervention, particularly among those with no prior pregnancy history.

The placenta is a dynamic organ that undergoes phases of development depending on the gestational age of the pregnancy.^9,10^ Placental biomarkers are shed into maternal circulation across gestation, with elevated or reduced concentrations of specific biomarkers in pregnant individuals who ultimately develop APOs.^11–14^ Biomarkers of interest include pregnancy-associated plasma protein A (PAPP-A), vascular endothelial growth factor (VEGF), placental growth factor (PlGF), soluble FMS-like tyrosine kinase 1 (sFlt-1), and soluble endoglin (sEng). Limited evidence shows that components of CVH in pregnancy may be associated with placental biomarkers^15,16^, and further data are needed to understand which components may be most closely related to circulating concentrations in early and mid-pregnancy.

The American Heart Association has set scoring standards for key components of “ideal” CVH, including health behaviors (physical activity, diet, smoking, and sleep duration) and health factors (blood pressure, glucose, lipids, and body mass index [BMI]).^17,18^ Higher (more favorable) CVH scores in pregnancy are associated with lower risk of adverse outcomes.^19^ While each individual CVH component is associated with pregnancy outcomes^15,20–25^, data associating these components with placental biomarkers are scant.^15,16^ This limits our understanding of the mechanisms involved in the relationship between CVH and APOs.

The purpose of this study was to examine whether CVH parameters measured in the 1^st^ trimester were associated with circulating maternal concentrations of placental biomarkers in the 1^st^ or 2^nd^ trimesters of pregnancy. We hypothesized that more ideal composite and individual component CVH scores measured in the 1^st^ trimester would be associated with more favorable biomarker concentrations including higher PAPP-A, VEGF, and PlGF, and lower sFlt-1 and sEng in the 1^st^ and 2^nd^ trimesters.

## Methods

### Sample

A subset of participants from the Nulliparous Pregnancy Outcomes Study: Monitoring Mothers to Be (nuMoM2b) prospective cohort comprised the sample for this study. Full methods for the cohort have been published previously.^26^ Briefly, nulliparous participants pregnant with a singleton gestation were recruited in early pregnancy (6^0^-13^6^ weeks’ gestation) and assessed three times throughout pregnancy (baseline, 16^0^-21^6^ weeks’ gestation and 22^0^-29^6^ weeks’ gestation) and at delivery at eight clinical sites in the United States. Ethical approval was obtained at each clinical site and participants provided informed consent prior to data collection. Biomarker assays were conducted on a subset of 1^st^ and 2^nd^ trimester maternal serum samples drawing from individuals who delivered after 20 weeks of gestation and provided sufficient serum.^11^ All cases of APO (preterm birth, preeclampsia, stillbirth, and/or small-for-gestational-age birth), as well as a random sample of 12% of the remaining individuals with uncomplicated, term deliveries, were included in the subset.^11^

### Cardiovascular Health Score

Each component of the CVH score was assessed in the 1^st^ trimester at the baseline visit and assigned a score on a scale of 0-100 according to the Life’s Essential 8 scoring algorithm, in which higher scores indicate more favorable health.^18^ The full scoring algorithm can be found in **Table S1**.

Physical activity was assessed via a self-report questionnaire, in which participants identified frequency, duration, and type of activities over the past four weeks. Average minutes per week of moderate (3.0-5.9 metabolic equivalents of task [METs]) and vigorous (≥6.0 METs) leisure time physical activity were calculated based on corresponding MET values from the Compendium of Physical Activities^27^, and weekly duration of physical activity as it pertains to U.S. federal guidelines was calculated as minutes of moderate plus 2 x minutes of vigorous activity.^28^ CVH scores ranged from 0 (no weekly physical activity) to 100 (≥150 minutes/week) with intermediate scores for values between 0-150 minutes/week. Physical activity was additionally categorized into none (no weekly physical activity), insufficient (1-149 minutes/week), sufficient (150-299 minutes/week), and high (300+ minutes/week).

The Healthy Eating Index (HEI) score was used to assess diet quality and was determined using a modified Block Food Frequency Questionnaire^29,30^ with reference to the three-month period immediately preceding conception. CVH scores were based on percentiles of HEI scores in the full nuMoM2b sample; for example, 0 points corresponded to HEI <25^th^ percentile, while 100 points corresponded to HEI ≥95^th^ percentile.

Self-reported weekday and weekend hours of nightly sleep were used to determine sleep duration. Weighted averages were calculated to account for differences between weekday and weekend sleep duration, and CVH scores were assigned based on the U-shaped association between sleep duration and cardiovascular health (i.e., both low and high sleep durations are adverse).

Participants self-reported combustible tobacco use and exposure to secondhand smoke in the home. CVH scoring for smoking was adapted from the standard scoring algorithm (see **Table S1**). A score of 0 indicated current smoking during pregnancy, while a score of 100 indicated never smoking tobacco; intermediate scores were assigned for former smokers. 20 points were subtracted in cases of exposure to secondhand smoke in the home for all except current smokers.

Blood pressure, height, and weight were measured in person by research personnel using standardized techniques. Body mass index (BMI, kg/m^2^) was calculated from height and weight. Blood draws were obtained in a non-fasted state, so the lipid and glucose components of the composite CVH score were excluded from this analysis. The composite CVH score was derived by averaging the scores of the six available components: physical activity, diet, sleep, smoking, blood pressure, and BMI. Composite CVH scores were categorized into high CVH (80-100 points), moderate CVH (50-79 points), and poor CVH (<50 points) for analysis.^18^

### Placental blood analytes

Full methodology of the biomarker assays have been published previously^11^; in brief, blood serum samples collected at the study visits in the 1^st^ and 2^nd^ trimester were stored at -70° C prior to being assayed. Biomarker concentrations were measured using enzyme-linked immunosorbent assay (sEng), electrochemiluminescence assay (VEGF and sFlt-1), or lanthanide-based time resolved fluorometry (PlGF and PAPP-A). Concentrations below the limits of detection were assigned values equal to half the lower limit, while those above the upper limit were assigned a value equal to the upper limit.

### Covariates

Demographic information including age, race/ethnicity, educational attainment, income, and health insurance status were self-reported via questionnaire at the baseline visit. At the second visit, participants completed the Krieger Discrimination Scale^31^ to assess experiences with racial discrimination and the Connor-Davidson Resilience Scale^32^ to assess resilience.

### Statistical Analyses

Distributions of placental blood biomarkers were examined for normality and log-transformed if appropriate. Descriptive characteristics were compared among composite CVH categories using Chi-square tests for categorical variables and analysis of variance for continuous variables.

Associations between a) composite CVH score categories and b) individual component scores with each placental blood biomarker were examined using linear regression. First, a cross-sectional analysis examined the association between 1^st^ trimester CVH metrics and 1^st^ trimester biomarker concentrations. Next, a longitudinal analysis examined the association between 1^st^ trimester CVH metrics and 2^nd^ trimester biomarker concentrations. Scatterplot visualizations of the associations between physical activity scores and the biomarkers suggested a nonlinear association, so physical activity was analyzed categorically (none, 0 minutes/week; insufficient, 1-149 minutes/week; sufficient, 150-299 minutes/week; and high, ≥300 minutes/week). BMI, blood pressure, diet, sleep duration, and smoking were analyzed as continuous CVH component scores.

Sampling weights were added to all analyses to account for the oversampling of APOs in the placental analyte subsample. All cases of APOs with sufficient serum were assayed (n=1,349); therefore, a sampling weight of 1 was applied to each APO case. Of the remaining participants with full-term uncomplicated pregnancy outcomes and sufficient serum, 11.8% were selected at random for assays (n=839). Thus, a sample weight of 8.48 (equal to 1/0.118) was applied to all non-APO controls. This method is similar to those used in nationally-representative cohorts such as the National Health and Nutrition Examination Survey^33^ and other prospective pregnancy cohort studies.^34^

All linear regression models were adjusted for gestational age at the blood draw, and longitudinal analyses were additionally adjusted for 1^st^ trimester biomarker concentrations. These minimally adjusted models were further adjusted for demographic characteristics, BMI (blood pressure and health behavior component analyses only), perceived racial discrimination, and resilience in a backwards stepwise approach in which covariate associations with a p-value <0.2 were retained in the final model (See **Table S2** for covariates in each model). All analyses were conducted in Stata SE version 18 (StataCorp, College Station, TX) and significant associations were accepted at p<0.01 to account for multiple comparisons while limiting Type II error.

## Results

### Participants

Of the 10,038 nuMoM2b participants, 2,188 were included in this analysis (**Figure 1**). Mean weighted baseline age of the included sample was 26.9±5.8 years and mean weighted BMI was 26.9±6.8 kg/m^2^. Participants included in this analysis had similar weighted rates of APOs but had significantly lower weighted BMI and were more likely to report non-Hispanic White race, lower perceived racial discrimination, and use of commercial insurance than the full cohort (**Table S3**).

**Figure 1.**
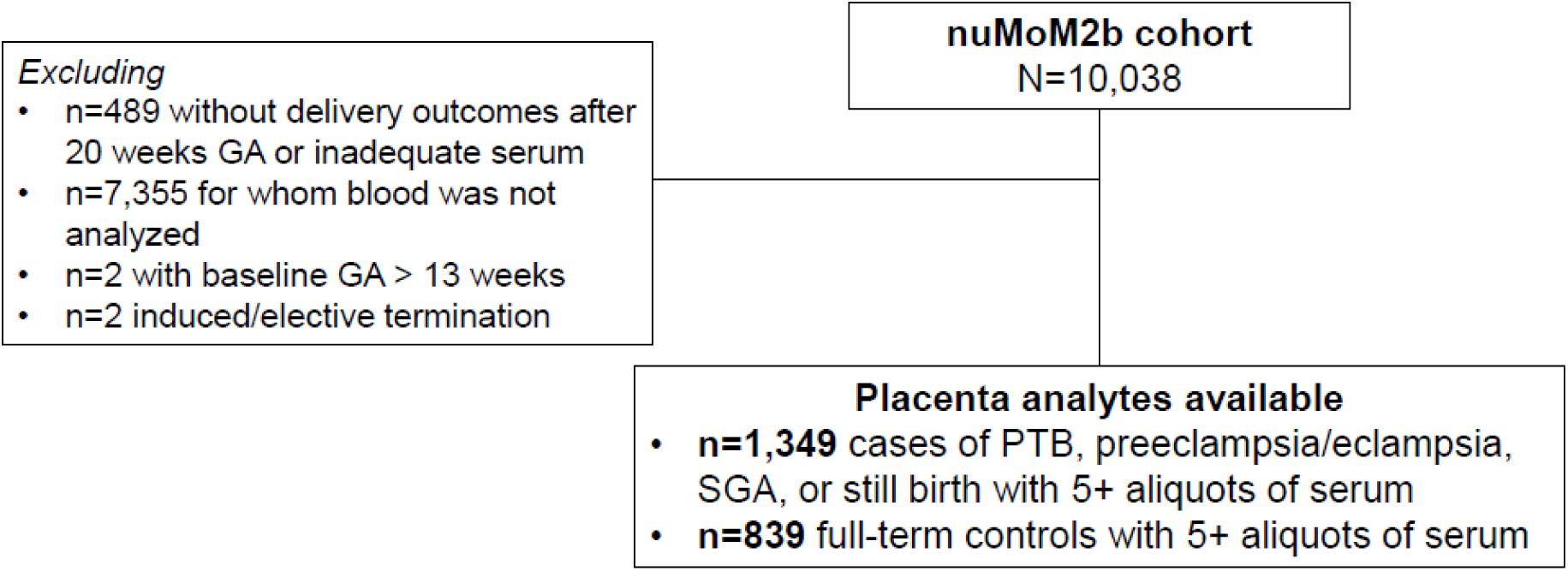
CONSORT diagram of included and excluded participants. nuMoM2b: Nulliparous Pregnancy Outcomes Study: Monitoring Mothers-to-be; GA: gestational age; SGA: small for gestational age; PTB: preterm (<37 weeks GA) birth.

### Composite CVH Scores

The mean weighted composite CVH score in the 1^st^ trimester for this sample was 71.1 ± 15.4 points. Thirty percent of participants achieved high CVH (≥80 points), 56% had moderate CVH (50-79 points), and 14% had poor CVH (<50 points, **Table 1**).^18^ Participants with high CVH were more likely to be older, non-Hispanic White, with a lower BMI and more education, higher income, or commercial insurance than those with moderate or poor CVH. Furthermore, the rate of APO was significantly lower in the high CVH group. Absolute concentrations of each placental biomarker by CVH group in each trimester can be found in **Table S4**.

**Table 1.** Descriptive characteristics of participants by physical activity categories. Results presented as weighted mean ± SD or frequency (weighted percentage) as appropriate.

|  | High CVH<br>(n=495) | Moderate CVH<br>(n=916) | Poor CVH<br>(n=220) | p-value |
| --- | --- | --- | --- | --- |
| **Maternal age (years)** | 28.8 ± 4.6 | 27.0 ± 5.8 | 24.9 ± 5.4 | **<0.001** |
| **BMI (kg/m<sup>2</sup>)** | 23.1 ± 3.1 | 26.4 ± 5.6 | 33.8 ± 9.4 | **<0.001** |
| **Maternal race** |  |  |  | **<0.001** |
| Asian | 29 (4%) | 23 (3%) | 1 (1%) |  |
| Black non-Hispanic | 17 (4%) | 119 (10%) | 69 (27%) |  |
| Hispanic | 50 (10%) | 182 (20%) | 37 (21%) |  |
| White non-Hispanic | 378 (79%) | 543 (63%) | 93 (44%) |  |
| Other | 21 (3%) | 49 (4%) | 20 (7%) |  |
| **Educational Attainment** |  |  |  | **<0.001** |
| Less than high school | 9 (1%) | 89 (10%) | 39 (14%) |  |
| Completed high school or GED | 14 (4%) | 129 (13%) | 57 (34%) |  |
| Some college | 45 (10%) | 190 (18%) | 69 (30%) |  |
| Associate or technical degree | 38 (9%) | 109 (9%) | 25 (9%) |  |
| Completed college | 185 (33%) | 239 (31%) | 18 (9%) |  |
| Degree work beyond college | 204 (43%) | 160 (19%) | 12 (4%) |  |
| **Percentage of Federal Poverty Level** |  |  |  | **<0.001** |
| <100% | 25 (5%) | 114 (15%) | 48 (40%) |  |
| 100-200% | 29 (8%) | 115 (12%) | 35 (28%) |  |
| >200% | 396 (87%) | 486 (73%) | 60 (32%) |  |
| **Insurance payor** |  |  |  | **<0.001** |
| Commercial | 331 (68%) | 472 (56%) | 88 (47%) |  |
| Medicare/Medicaid | 25 (5%) | 272 (26%) | 107 (43%) |  |
| Military/Other | 2 (1%) | 14 (1%) | 3 (<1%) |  |
| Two or more sources | 125 (23%) | 137 (16%) | 11 (7%) |  |
| None (personal income) | 11 (3%) | 10 (1%) | 6 (3%) |  |
| **Perceived racial discrimination** |  |  |  | **0.001** |
| None | 398 (81%) | 665 (74%) | 148 (68%) |  |
| Moderate | 67 (14%) | 180 (19%) | 46 (18%) |  |
| High | 30 (5%) | 71 (7%) | 26 (14%) |  |
| **Resilience** | 82.5 ± 10.1 | 78.4 ± 12.3 | 77.5 ± 11.6 | **<0.001** |
| **Composite CVH score** | 87.3 ± 5.3 | 66.3 ± 8.3 | 42.5 ± 6.6 | **<0.001** |
| **APO** | 268 (13%) | 553 (16%) | 164 (27%) | **<0.001** |

*High cardiovascular health (CVH): 80-100 points; Moderate CVH: 50-79 points; Poor CVH: <50 points. Significance accepted at $p < 0.01$ . Perceived racial discrimination scores from Krieger discrimination scale: None: 0; Moderate: 1-2; High: 3+. Resilience scores on a scale from 0-100. APO: adverse pregnancy outcome; BMI: body mass index; GED: general education diploma.*

### Circulating PAPP-A

High CVH was associated with a higher (more favorable) adjusted 1^st^ trimester circulating PAPP-A concentration (2.98 log mU/mL, 95% CI [2.93, 3.04]) than both moderate CVH (2.88 log mU/mL, 95% CI [2.82, 2.92], p=0.004) and poor CVH (2.61 log mU/mL, 95% CI [2.49, 2.72], p<0.001). Results were similar when evaluated in longitudinal analysis estimating the association of 1^st^ trimester composite CVH with 2^nd^ trimester PAPP-A concentrations (3.94 log mU/mL, 95% CI [3.91, 3.96] for those with high CVH versus 3.82 log mU/ml, 95% CI [3.75, 3.89] for those with poor CVH, p=0.002).

### Circulating VEGF, PlGF, sFlt-1, and sEng

The high vs. poor CVH group had lower adjusted circulating VEGF in the 1^st^ trimester (-0.13 log pg/mL, 95% CI [-0.18, -0.08] versus 0.10 log pg/mL, 95% CI [-0.05, 0.24], p=0.006) and higher circulating sFlt-1 concentrations in the 1^st^ trimester (2.97 log pg/mL, 95% CI [2.95, 2.99] versus 2.90 log pg/mL, 95% CI [2.86, 2.94], p=0.004; **Table 2**). Furthermore, high CVH was associated with higher adjusted circulating 1^st^ trimester sEng than poor CVH (6.81 ng/mL, 95% CI [6.61, 7.00] versus 5.79 ng/mL, 95% CI [5.43, 6.15], p<0.001). No longitudinal associations were detected between composite CVH and 2^nd^ trimester angiogenic biomarker concentrations.

**Table 2.**
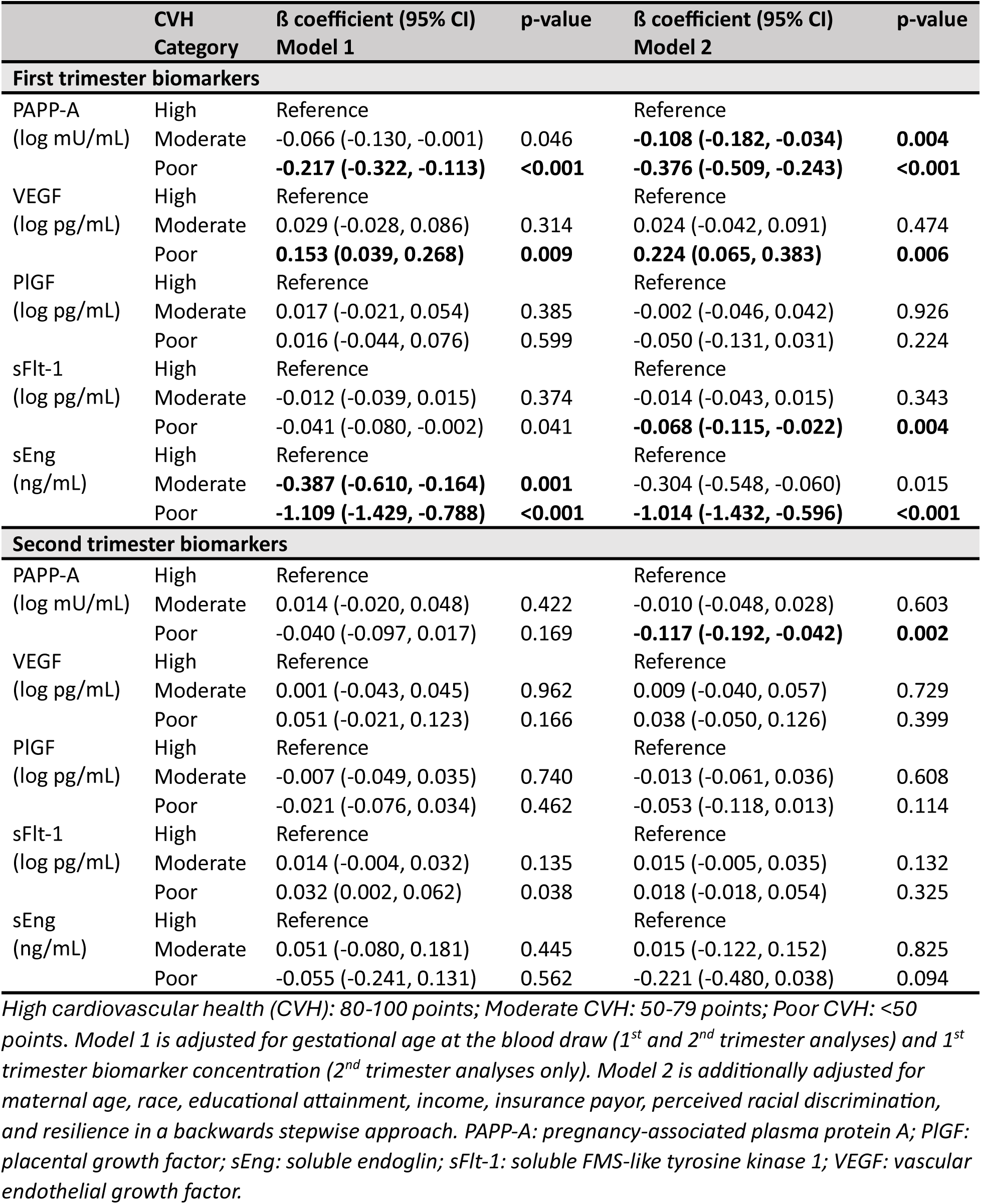
Association between 1^st^ trimester cardiovascular health score categories and placental biomarkers in the 1^st^ and 2^nd^ trimesters.

| | CVH Category | $\beta$ coefficient (95% CI)<br>Model 1 | p-value | $\beta$ coefficient (95% CI)<br>Model 2 | p-value |
| --- | --- | --- | --- | --- | --- |
| **First trimester biomarkers** |  |  |  |  |  |
| PAPP-A | High | Reference |  | Reference |  |
| (log mU/mL) | Moderate | -0.066 (-0.130, -0.001) | 0.046 | **-0.108 (-0.182, -0.034)** | **0.004** |
|  | Poor | **-0.217 (-0.322, -0.113)** | **<0.001** | **-0.376 (-0.509, -0.243)** | **<0.001** |
| VEGF | High | Reference |  | Reference |  |
| (log pg/mL) | Moderate | 0.029 (-0.028, 0.086) | 0.314 | 0.024 (-0.042, 0.091) | 0.474 |
|  | Poor | **0.153 (0.039, 0.268)** | **0.009** | **0.224 (0.065, 0.383)** | **0.006** |
| PlGF | High | Reference |  | Reference |  |
| (log pg/mL) | Moderate | 0.017 (-0.021, 0.054) | 0.385 | -0.002 (-0.046, 0.042) | 0.926 |
|  | Poor | 0.016 (-0.044, 0.076) | 0.599 | -0.050 (-0.131, 0.031) | 0.224 |
| sFlt-1 | High | Reference |  | Reference |  |
| (log pg/mL) | Moderate | -0.012 (-0.039, 0.015) | 0.374 | -0.014 (-0.043, 0.015) | 0.343 |
|  | Poor | -0.041 (-0.080, -0.002) | 0.041 | **-0.068 (-0.115, -0.022)** | **0.004** |
| sEng | High | Reference |  | Reference |  |
| (ng/mL) | Moderate | **-0.387 (-0.610, -0.164)** | **0.001** | -0.304 (-0.548, -0.060) | 0.015 |
|  | Poor | **-1.109 (-1.429, -0.788)** | **<0.001** | **-1.014 (-1.432, -0.596)** | **<0.001** |
| **Second trimester biomarkers** |  |  |  |  |  |
| PAPP-A | High | Reference |  | Reference |  |
| (log mU/mL) | Moderate | 0.014 (-0.020, 0.048) | 0.422 | -0.010 (-0.048, 0.028) | 0.603 |
|  | Poor | -0.040 (-0.097, 0.017) | 0.169 | **-0.117 (-0.192, -0.042)** | **0.002** |
| VEGF | High | Reference |  | Reference |  |
| (log pg/mL) | Moderate | 0.001 (-0.043, 0.045) | 0.962 | 0.009 (-0.040, 0.057) | 0.729 |
|  | Poor | 0.051 (-0.021, 0.123) | 0.166 | 0.038 (-0.050, 0.126) | 0.399 |
| PlGF | High | Reference |  | Reference |  |
| (log pg/mL) | Moderate | -0.007 (-0.049, 0.035) | 0.740 | -0.013 (-0.061, 0.036) | 0.608 |
|  | Poor | -0.021 (-0.076, 0.034) | 0.462 | -0.053 (-0.118, 0.013) | 0.114 |
| sFlt-1 | High | Reference |  | Reference |  |
| (log pg/mL) | Moderate | 0.014 (-0.004, 0.032) | 0.135 | 0.015 (-0.005, 0.035) | 0.132 |
|  | Poor | 0.032 (0.002, 0.062) | 0.038 | 0.018 (-0.018, 0.054) | 0.325 |
| sEng | High | Reference |  | Reference |  |
| (ng/mL) | Moderate | 0.051 (-0.080, 0.181) | 0.445 | 0.015 (-0.122, 0.152) | 0.825 |
|  | Poor | -0.055 (-0.241, 0.131) | 0.562 | -0.221 (-0.480, 0.038) | 0.094 |

High cardiovascular health (CVH): 80-100 points; Moderate CVH: 50-79 points; Poor CVH: <50 points. Model 1 is adjusted for gestational age at the blood draw (1<sup>st</sup> and 2<sup>nd</sup> trimester analyses) and 1<sup>st</sup> trimester biomarker concentration (2<sup>nd</sup> trimester analyses only). Model 2 is additionally adjusted for maternal age, race, educational attainment, income, insurance payor, perceived racial discrimination, and resilience in a backwards stepwise approach. PAPP-A: pregnancy-associated plasma protein A; PlGF: placental growth factor; sEng: soluble endoglin; sFlt-1: soluble FMS-like tyrosine kinase 1; VEGF: vascular endothelial growth factor.

### Individual Component Scores

#### Circulating PAPP-A

Higher (more favorable) component scores for BMI and blood pressure were associated with higher 1^st^ trimester PAPP-A concentrations in minimally-adjusted models (**Table 3**, p<0.001 for each), but only BMI remained significantly associated following covariate adjustment. Similar results were detected when BMI was evaluated longitudinally with PAPP-A concentrations in the 2^nd^ trimester. No significant associations were found between component scores for any of the health behaviors and 1^st^ trimester PAPP-A concentrations.

**Table 3.** Association between 1^st^ trimester component scores and placental biomarkers in the 1^st^ and 2^nd^ trimesters. ß coefficients are per 10-point increase in component score. A higher score is healthier.

| | $\beta$ coefficient per 10-point increase in CVH score (95% CI)<br>Model 1 | p-value | $\beta$ coefficient per 10-point increase in CVH score (95% CI)<br>Model 2 | p-value |
| --- | --- | --- | --- | --- |
| **First trimester biomarkers** |  |  |  |  |
| **Log PAPP-A (mU/mL)** |  |  |  |  |
| Body Mass Index | **0.046 (0.038, 0.054)** | **<0.001** | **0.056 (0.046, 0.066)** | **<0.001** |
| Blood Pressure | **0.050 (0.032, 0.065)** | **<0.001** | 0.022 (0.002, 0.043) | 0.033 |
| Smoking | -0.0002 (-0.008, 0.008) | 0.956 | -0.0002 (-0.010, 0.009) | 0.964 |
| Diet | -0.003 (-0.011, 0.006) | 0.561 | -0.004 (-0.015, 0.008) | 0.506 |
| Sleep | 0.002 (-0.012, 0.016) | 0.788 | 0.005 (-0.011, 0.021) | 0.559 |
| **Log VEGF (pg/mL)** |  |  |  |  |
| Body Mass Index | **-0.040 (-0.050, -0.031)** | **<0.001** | **-0.044 (-0.054, -0.033)** | **<0.001** |
| Blood Pressure | **-0.035 (-0.050, -0.020)** | **<0.001** | -0.011 (-0.027, 0.006) | 0.196 |
| Smoking | -0.004 (-0.011, 0.003) | 0.280 | 0.002 (-0.006, 0.009) | 0.668 |
| Diet | -0.0003 (-0.008, 0.008) | 0.933 | 0.008 (-0.002, 0.017) | 0.118 |
| Sleep | -0.003 (-0.016, 0.009) | 0.597 | -0.007 (-0.022, 0.008) | 0.341 |
| **Log PIGF (pg/mL)** |  |  |  |  |
| Body Mass Index | 0.002 (-0.002, 0.006) | 0.281 | 0.003 (-0.002, 0.008) | 0.262 |
| Blood Pressure | 0.006 (-0.001, 0.012) | 0.072 | 0.006 (-0.001, 0.013) | 0.094 |
| Smoking | **-0.006 (-0.010, -0.002)** | **0.004** | -0.006 (-0.010, -0.0001) | 0.018 |
| Diet | -0.005 (-0.009, -0.0001) | 0.046 | 0.001 (-0.005, 0.007) | 0.772 |
| Sleep | -0.002 (-0.008, 0.004) | 0.559 | 0.004 (-0.003, 0.011) | 0.317 |
| **Log sFlt-1 (pg/mL)** |  |  |  |  |
| Body Mass Index | **0.014 (0.010, 0.017)** | **<0.001** | **0.015 (0.011, 0.019)** | **<0.001** |
| Blood Pressure | **0.012 (0.006, 0.017)** | **<0.001** | 0.002 (-0.005, 0.008) | 0.592 |
| Smoking | 0.001 (-0.002, 0.004) | 0.392 | -0.0004 (-0.004, 0.003) | 0.837 |
| Diet | -0.001 (-0.004, 0.003) | 0.729 | -0.003 (-0.007, 0.001) | 0.188 |
| Sleep | -0.002 (-0.007, 0.004) | 0.506 | 0.001 (-0.006, 0.009) | 0.691 |
| **sEng (ng/mL)** |  |  |  |  |
| Body Mass Index | **0.166 (0.142, 0.190)** | **<0.001** | **0.164 (0.137, 0.192)** | **<0.001** |
| Blood Pressure | **0.113 (0.070, 0.156)** | **<0.001** | -0.031 (-0.079, 0.018) | 0.212 |
| Smoking | **0.050 (0.024, 0.076)** | **<0.001** | 0.035 (0.008, 0.062) | 0.011 |
| Diet | 0.034 (0.004, 0.063) | 0.026 | 0.014 (-0.019, 0.046) | 0.416 |
| Sleep | **0.064 (0.020, 0.11)** | **0.004** | 0.046 (-0.005, 0.097) | 0.075 |
| **Second trimester biomarkers** |  |  |  |  |
| **Log PAPP-A (mU/mL)** |  |  |  |  |
| Body Mass Index | **0.017 (0.011, 0.023)** | **<0.001** | **0.024 (0.018, 0.031)** | **<0.001** |
| Blood Pressure | 0.010 (0.001, 0.018) | 0.023 | 0.001 (-0.008, 0.010) | 0.846 |
| Smoking | 0.005 (0.0001, 0.009) | 0.044 | 0.004 (-0.001, 0.009) | 0.088 |
| Diet | -0.003 (-0.008, 0.002) | 0.284 | -0.002 (-0.007, 0.004) | 0.561 |
| Sleep | -0.005 (-0.012, 0.003) | 0.243 | -0.004 (-0.013, 0.006) | 0.442 |

| **Log VEGF (pg/mL)** |  |  |  |  |
| --- | --- | --- | --- | --- |
| Body Mass Index | -0.004 (-0.010, 0.001) | 0.117 | -0.006 (-0.013, 0.0002) | 0.059 |
| Blood Pressure | -0.004 (-0.012, 0.005) | 0.412 | -0.003 (-0.014, 0.007) | 0.525 |
| Smoking | -0.0004 (-0.006, 0.005) | 0.877 | 0.0006 (-0.006, 0.007) | 0.849 |
| Diet | -0.002 (-0.008, 0.004) | 0.556 | -0.0001 (-0.007, 0.007) | 0.975 |
| Sleep | 0.002 (-0.007, 0.011) | 0.691 | 0.006 (-0.006, 0.017) | 0.351 |
| **Log PlGF (pg/mL)** |  |  |  |  |
| Body Mass Index | **0.013 (0.009, 0.017)** | **<0.001** | **0.013 (0.008, 0.018)** | **<0.001** |
| Blood Pressure | 0.012 (0.002, 0.023) | 0.025 | 0.005 (-0.008, 0.017) | 0.492 |
| Smoking | 0.0004 (-0.004, 0.005) | 0.858 | 0.003 (-0.003, 0.009) | 0.323 |
| Diet | 0.0007 (-0.006, 0.007) | 0.828 | -0.002 (-0.001, 0.007) | 0.732 |
| Sleep | -0.003 (-0.009, 0.004) | 0.427 | -0.0001 (-0.008, 0.007) | 0.983 |
| **Log sFlt-1 (pg/mL)** |  |  |  |  |
| Body Mass Index | 0.0001 (-0.003, 0.003) | 0.953 | 0.001 (-0.002, 0.004) | 0.431 |
| Blood Pressure | -0.004 (-0.008, 0.0002) | 0.065 | -0.001 (-0.005, 0.003) | 0.746 |
| Smoking | 0.0002 (-0.002, 0.003) | 0.860 | 0.001 (-0.002, 0.004) | 0.430 |
| Diet | -0.001 (-0.004, 0.001) | 0.347 | 0.0003 (-0.003, 0.004) | 0.849 |
| Sleep | -0.0004 (-0.004, 0.004) | 0.853 | -0.001 (-0.006, 0.004) | 0.823 |
| **sEng (ng/mL)** |  |  |  |  |
| Body Mass Index | 0.004 (-0.011, 0.020) | 0.582 | 0.006 (-0.013, 0.025) | 0.515 |
| Blood Pressure | -0.011 (-0.037, 0.014) | 0.377 | -0.012 (-0.044, 0.019) | 0.442 |
| Smoking | -0.007 (-0.022, 0.008) | 0.362 | 0.005 (-0.013, 0.023) | 0.590 |
| Diet | -0.004 (-0.022, 0.014) | 0.658 | -0.001 (-0.025, 0.023) | 0.917 |
| Sleep | -0.011 (-0.039, 0.017) | 0.441 | -0.007 (-0.038, 0.024) | 0.659 |

*Model 1 is adjusted for gestational age at the blood draw (1<sup>st</sup> and 2<sup>nd</sup> trimester analyses) and 1<sup>st</sup> trimester biomarker concentration (2<sup>nd</sup> trimester analyses only). Model 2 is additionally adjusted for maternal age, race, body mass index, educational attainment, income, insurance payor, perceived racial discrimination, and resilience in a backwards stepwise approach. PAPP-A: pregnancy-associated plasma protein A; PlGF: placental growth factor; sEng: soluble endoglin; sFlt-1: soluble FMS-like tyrosine kinase 1; VEGF: vascular endothelial growth factor.*

Participants reporting no weekly 1^st^ trimester physical activity had significantly higher 2^nd^ trimester PAPP-A than those reporting insufficient, sufficient, or high physical activity (**Figure 2a**, p<0.001 for all). While 1^st^ trimester physical activity was associated with lower 2^nd^ trimester concentrations of PAPP-A, a significantly lower proportion of those reporting sufficient weekly physical activity had an APO compared to those reporting no weekly physical activity (**Figure 2b**, p<0.01).

**Figure 2.**
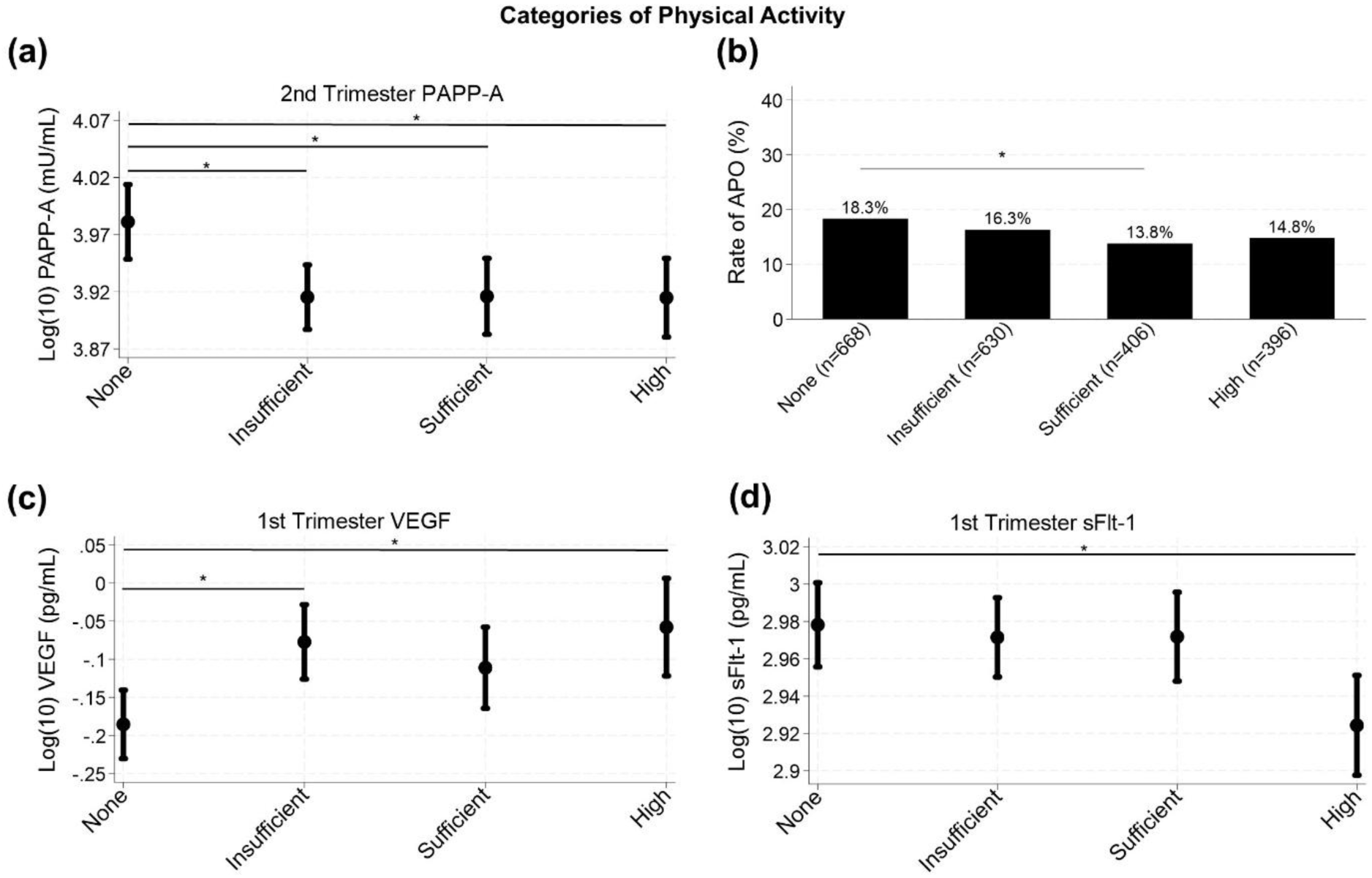
Differences between physical activity categories, defined as: None = 0 minutes/week; Insufficient = 1-149 minutes per week; Sufficient = 150-299 minutes/week; High = ≥300 minutes/week. *Indicates p<0.01 **(a)** Any 1^st^ trimester physical activity associated with lower 2^nd^ trimester PAPP-A than none; **(b)** Significantly lower weighted rate of APO in those with sufficient weekly physical activity compared to none; **(c)** No weekly 1^st^ trimester physical activity associated with lower 1^st^ trimester VEGF than those with insufficient or high weekly physical activity (p<0.001), while those with sufficient weekly physical activity were non-significantly lower (p=0.02); **(d)** High weekly 1^st^ trimester physical activity associated with lower 1^st^ trimester sFlt-1 than no weekly physical activity. PAPP-A: pregnancy-associated plasma protein A; sFlt-1: soluble FMS-like tyrosine kinase 1; VEGF: vascular endothelial growth factor. Data presented as weighted means with 95% confidence intervals **(a, c, d)** or weighted percentages **(b)**.

#### Circulating VEGF, PlGF, sFlt-1, and sEng

Higher (more favorable) component scores for BMI and blood pressure were significantly associated with lower 1^st^ trimester concentrations of VEGF, higher 1^st^ trimester concentrations of sFlt-1, and higher 1^st^ trimester concentrations of sEng; only the associations with BMI remained significant following covariate adjustment (**Table 3**). In longitudinal analyses, a more favorable 1^st^ trimester BMI score was associated with a higher 2^nd^ trimester concentration of PlGF, which was robust to covariate adjustment.

Engaging in physical activity was associated with a higher 1^st^ trimester VEGF concentration (**Figure 2c**, p<0.001 for insufficient and high physical activity compared to none). High physical activity was associated with lower 1^st^ trimester sFlt-1 concentration versus no weekly physical activity (**Figure 2d**, p=0.003). These associations with physical activity were robust to covariate adjustment. No associations with blood pressure, smoking, diet, or sleep scores were detected following covariate adjustment (**Tables 3 & S5**).

## Discussion

Our findings demonstrate the complex interplay between maternal CVH and placental biomarkers in nulliparous pregnant participants, with associations in both expected and unexpected directions (**Figure 3**). BMI emerged as the only component that was significantly associated with each circulating biomarker (PAPP-A, VEGF, sFlt-1, and sEng in the 1^st^ trimester, and PAPP-A and PlGF in the 2^nd^ trimester), though associations with 1^st^ trimester angiogenic biomarkers were in the opposite direction as hypothesized.

**Figure 3.**
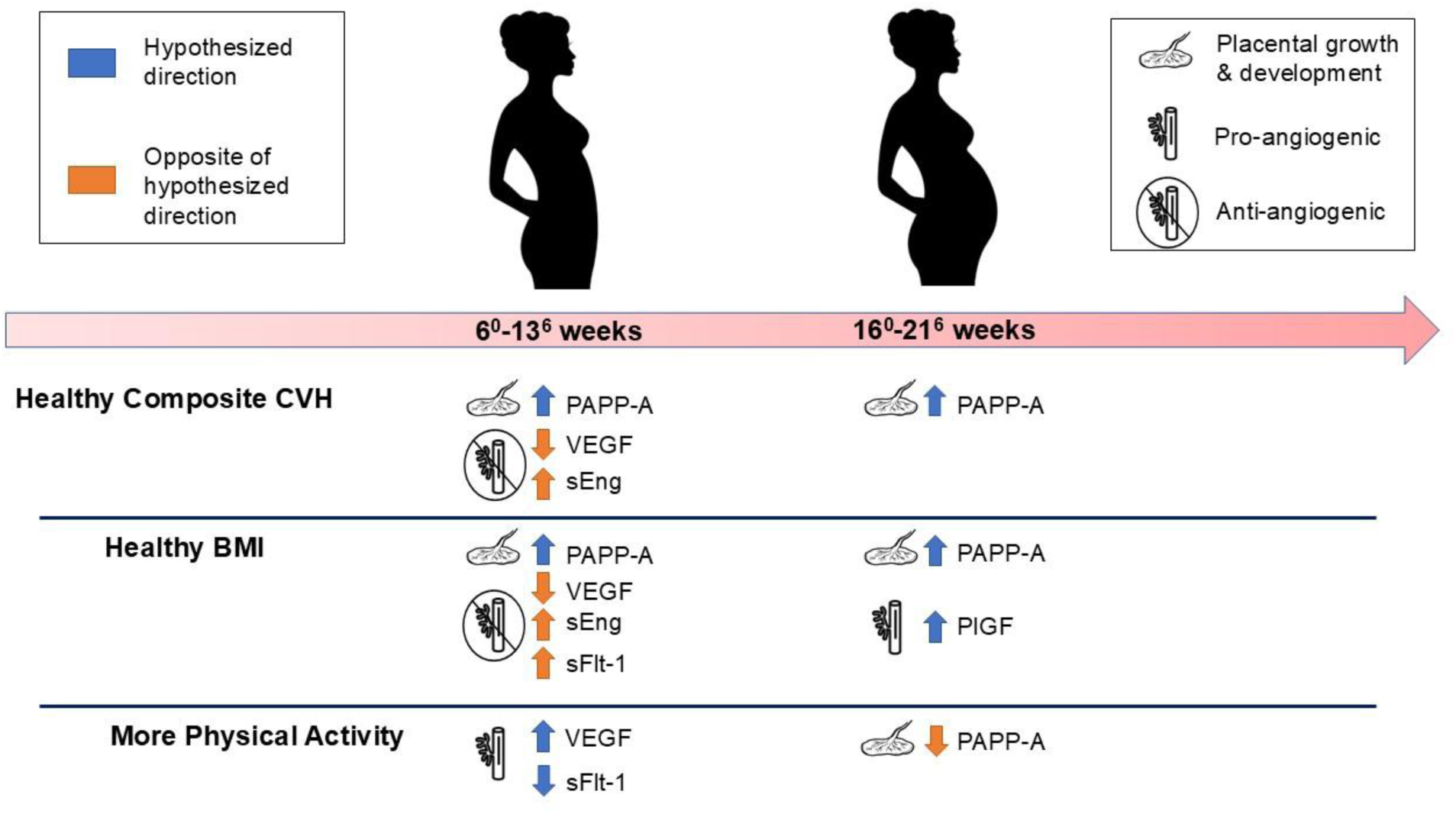
Associations of composite cardiovascular health (CVH), BMI, and physical activity components with circulating placental biomarker concentrations in the 1^st^ and 2^nd^ trimesters. Hypothesis: healthier CVH and components are associated with higher PAPP-A, VEGF, and PlGF, and with lower sEng and sFlt-1. PAPP-A: pregnancy-associated plasma protein A; sFlt-1: soluble FMS-like tyrosine kinase 1; VEGF: vascular endothelial growth factor.

Physical activity was the only health behavior that was associated with biomarkers independent of BMI, with expected directions of associations with 1^st^ trimester angiogenic biomarkers and an unexpected direction with 2^nd^ trimester PAPP-A. These results suggest that BMI is an important factor relating to circulating placental biomarkers in early and mid-pregnancy, and physical activity may be the predominant health behavior relating to placental biomarkers during this first half of pregnancy.

### Circulating PAPP-A

PAPP-A concentrations increase across gestation in normal pregnancy^35,36^ and display dysregulated trajectories in cases of APOs.^37^ In preeclampsia, 1^st^ trimester PAPP-A is lower than normal, then increases exponentially in the 3^rd^ trimester to levels above normal.^14^ We found that those with favorable 1^st^ trimester CVH had higher 1^st^ trimester circulating PAPP-A than those with moderate or poor CVH. As higher concentrations of this biomarker in early gestation are associated with better pregnancy outcomes, this finding suggests that higher early-pregnancy CVH may be related to more robust placentation and concomitant higher concentrations of PAPP-A. Furthermore, the strong association between more favorable BMI component scores and higher PAPP-A suggests that a healthy BMI, in particular, may be important for healthy placentation.

In contrast, the association between greater 1^st^ trimester physical activity and lower 2^nd^ trimester concentrations of PAPP-A suggests that physical activity in early pregnancy may be related to less robust placental growth and development by mid-pregnancy. This finding is in the opposite direction from our hypothesis, yet it is important to note that it did not appear to be pathologic – participants who self-reported at least 150 minutes/week of 1^st^ trimester physical activity had a significantly lower rate of APOs than those who self-reported no weekly physical activity. We speculate this finding may be due to the timing of the PAPP-A measurement. In the 2^nd^ trimester, PAPP-A concentrations are similar between individuals who ultimately develop healthy and adverse pregnancy outcomes; low PAPP-A is adverse in the 1^st^ trimester, but high PAPP-A is adverse in the 3^rd^ trimester.^12,14^ It is possible that the lower 2^nd^ trimester PAPP-A concentrations we found in those engaging in physical activity may be indicative of physical activity eliciting a protective effect against higher late-pregnancy concentrations, but this warrants further study.

### Circulating VEGF, PlGF, sFlt-1, and sEng

We found that healthier 1^st^ trimester composite CVH scores and BMI component scores were associated with lower concentrations of VEGF, while more self-reported 1^st^ trimester physical activity was associated with higher 1^st^ trimester VEGF. Outside of pregnancy, higher VEGF concentrations are associated with obesity and metabolic syndrome^38–40^ which may be due to the angiogenic burden to support adipose tissue. However, exercise is also associated with higher VEGF expression and circulating concentrations outside pregnancy, though from a physiological perspective of supporting oxygen delivery to the musculature.^40–42^ Limited pregnancy literature supports both these associations, with higher placental VEGF expression in those with gestational diabetes and high gestational weight gain^43,44^ as well as in active versus inactive pregnant individuals.^45^ It is important to note that these angiogenic biomarkers are produced by several tissue types in addition to the placenta, and circulating concentrations cannot discriminate between placental versus maternal tissue origins. It is possible that the associations we detected are indicative of both placental and maternal tissue contributions to circulating VEGF concentrations.

Similarly, we found that healthier 1^st^ trimester composite CVH scores and BMI component scores were associated with higher concentrations of sEng, opposite to our hypothesis. Higher concentrations of sEng in early- and mid-pregnancy are associated with higher risk for preeclampsia.^13,46,47^ However, maternal obesity has been linked with lower circulating concentrations of sEng.^48^ We also found healthier BMI component scores were associated with higher 1^st^ trimester sFlt-1. This finding supports prior literature demonstrating that maternal obesity is associated with lower sFlt-1 in early pregnancy, which may be related to greater plasma volume or higher concentrations of sFlt-1 degrading heparin sulfate proteoglycans.^48–51^ Healthier BMI scores were also associated with higher 2^nd^ trimester PlGF concentrations in our study. Prior literature has found similar results, with lower mid-pregnancy PlGF concentrations found in those with higher BMI.^48,51^ It is possible that a healthy early-pregnancy BMI is related to a more robust placenta that is capable of synthesizing sufficient PlGF by mid-pregnancy.

Finally, we found that more self-reported 1^st^ trimester physical activity was associated with lower sFlt-1 concentrations in the 1^st^ trimester. This confirms the findings of a prior study in the same cohort that only examined physical activity, and found significantly lower sFlt-1 concentrations in those above versus below 150 min/week physical activity.^15^ Our results expand on this finding, as we demonstrated that the lowest concentration of sFlt-1 was seen in those reporting ≥300 min/week physical activity and that those reporting 150-299 min/week were not significantly different from those reporting no weekly physical activity. This suggests that physical activity may indeed be a potential mechanism to reduce the antiangiogenic sFlt-1 concentration in early pregnancy^15^, but it may require high volumes of physical activity to reach this target.

### Strengths and Limitations

This study is bolstered by several strengths. We used a diverse national cohort of nulliparous individuals, in which robust data were collected across pregnancy. This allowed us to examine both cross-sectional and longitudinal associations between CVH and placental biomarkers in the 1^st^ and 2^nd^ trimesters, which is of particular importance given the dynamic nature of placental development throughout pregnancy.^52,53^

Certain limitations are acknowledged. The sample for this analysis was comprised of all cases of APO and a random sample of remaining participants with uncomplicated full-term pregnancies from the parent study. While we used sampling weights to account for this, some differences remained between our weighted sample and the full cohort. Multiple comparisons may have led to a Type I error, though we used an *a priori* cutoff of p<0.01 to mitigate this risk. By only analyzing those whose pregnancies continued beyond 20 weeks of gestation our results may be impacted by survivorship bias. Health behaviors were all self-reported, although participants were not aware of biomarker concentrations at the time of behavior recall so non-random bias should be minimal. The diet assessment referenced the 3 months prior to conception and may not have been representative of 1^st^ trimester dietary intake. We did not have fasting blood glucose or lipids and so were not able to examine the full spectrum of CVH components, and we did not have placental histology data to directly examine morphology outcomes. Finally, we did not adjust for medications; while it is possible that aspirin therapy could have impacted our results, prior analysis in this cohort found less than 2% of participants reported aspirin use during their pregnancy and a recent clinical trial found no effect of aspirin therapy on circulating concentrations of PAPP-A and PlGF.^54,55^

### Conclusions

Maternal CVH in the 1^st^ trimester of pregnancy is associated with circulating concentrations of placental biomarkers. While many of our findings were in the hypothesized direction wherein a healthier CVH score was associated with healthier circulating concentrations, others were in the opposite direction. Our findings suggest that maternal BMI may have a complex relationship with these biomarkers. In some cases (PAPP-A in both trimesters and 2^nd^ trimester PlGF), a healthier BMI was indicative of more robust placentation. In contrast, a less favorable BMI was associated with a more pro-angiogenic balance of 1^st^ trimester VEGF, sFlt-1, and sEng. We cautiously interpret this finding to suggest that maternal systemic factors such as the higher angiogenic demand of adipose tissue in the setting of obesity may play a larger role than placental factors in angiogenic protein concentrations in early pregnancy. Finally, of the five remaining CVH components (blood pressure, physical activity, diet, sleep duration, and smoking), only physical activity was independently associated with circulating biomarker concentrations when accounting for BMI. Future investigations of biomarker concentrations in early pregnancy should include BMI as a covariate, and physical activity may be a target for interventions in this population.

## Data Availability

Publicly-available data related to this study are available on the DASH data and specimen hub.

## Non-standard Abbreviations and Acronyms

APO: Adverse pregnancy outcomes
BMI: Body mass index
CVH: Cardiovascular health
HEI: Healthy Eating Index
METs: Metabolic equivalents of the task
nuMoM2b: Nulliparous Pregnancy Outcomes Study: Monitoring Mothers-to-be
PAPP-A: Pregnancy-associated plasma protein A
PlGF: Placental growth factor
PTB: Preterm birth
sEng: Soluble endoglin
sFlt-1: Soluble FMS-like tyrosine kinase 1
SGA: Small for gestational age
VEGF: Vascular endothelial growth factor

## Supplemental Tables

**Table S1.** Cardiovascular Health Scoring algorithm for health behaviors in pregnancy. HEI: Healthy Eating Index.

|  | Value | Cardiovascular Health Score |
| --- | --- | --- |
| Physical Activity<br>(minutes moderate-<br>to-vigorous physical<br>activity/week) | 0 | 0 |
|  | 1-29 | 20 |
|  | 30-59 | 40 |
|  | 60-89 | 60 |
|  | 90-119 | 80 |
|  | 120-149 | 90 |
|  | 150+ | 100 |
| Diet (HEI percentile) | <25<sup>th</sup> | 0 |
|  | 25<sup>th</sup>-49<sup>th</sup> | 25 |
|  | 50<sup>th</sup>-74<sup>th</sup> | 50 |
|  | 75<sup>th</sup>-94<sup>th</sup> | 80 |
|  | ≥95<sup>th</sup> | 100 |
| Sleep Duration (mean<br>hours/night) | <4 | 0 |
|  | 4 - <5 | 20 |
|  | 5 - <6 or ≥10 | 40 |
|  | 6 - <7 | 70 |
|  | 9 - <10 | 90 |
|  | 7 - <9 | 100 |
| Smoking | Current smoking | 0 |
|  | Smoked within 3 months of<br>conception | 25 |
|  | Former smoker, quit prior<br>to 3 months before<br>conception | 50 |
|  | Never smoker | 100 |
|  | Lives with current smoker | Subtract 20 points unless<br>score already = 0 |

**Table S2.** Covariates included in each model following backwards stepwise selection (p to remain <0.2).

|  | Cross-sectional analysis | Longitudinal analysis |
| --- | --- | --- |
| ***PAPP-A*** |  |  |
| Composite CVH Score | Age, education, race/ethnicity, insurance | Age, discrimination, income, race/ethnicity |
| BMI | Age, education, insurance, race/ethnicity | Race/ethnicity, income, resilience |
| Blood Pressure | Age, BMI, race/ethnicity, insurance | Race/ethnicity, BMI, income |
| Physical Activity | Age, BMI, discrimination, race/ethnicity, insurance | Race/ethnicity, BMI, income |
| Smoking | Age, BMI, education, race/ethnicity, insurance | Age, BMI, education, income, race/ethnicity |
| Diet | Age, BMI, education, race/ethnicity, insurance | Race/ethnicity, BMI, discrimination, income |
| Sleep | Age, BMI, education, race/ethnicity, insurance | Race/ethnicity, BMI, discrimination, income |
| ***VEGF*** |  |  |
| Composite CVH Score | Education | Race/ethnicity |
| BMI | Age, income | Race/ethnicity |
| Blood Pressure | Age, BMI, income | Race/ethnicity, BMI |
| Physical Activity | Age, BMI | Race/ethnicity, BMI |
| Smoking | Age, BMI, income | Race/ethnicity, BMI |
| Diet | Age, BMI, income | Race/ethnicity, BMI, discrimination |
| Sleep | Age, BMI, income | Race/ethnicity, BMI |
| ***PIGF*** |  |  |
| Composite CVH Score | Discrimination, education, income, race/ethnicity | Race/ethnicity |
| BMI | Race/ethnicity | Age, race/ethnicity |
| Blood Pressure | Race/ethnicity | Age, BMI, education, race/ethnicity |
| Physical Activity | Race/ethnicity, BMI, education | Age, BMI, race/ethnicity |
| Smoking | Race/ethnicity, BMI | Age, BMI, education, race/ethnicity |
| Diet | Discrimination, BMI, education, income, race/ethnicity | Age, BMI, race/ethnicity |
| Sleep | Discrimination, BMI, education, income, race/ethnicity | Age, BMI, race/ethnicity |
| ***sFlt-1*** |  |  |
| Composite CVH Score | Education, race/ethnicity, discrimination | Race/ethnicity |
| BMI | Insurance, race/ethnicity | Age, race/ethnicity, education, discrimination |
| Blood Pressure | Insurance, BMI, race/ethnicity | Race/ethnicity, education |
| Physical Activity | Insurance, BMI, race/ethnicity | Race/ethnicity, education |
| Smoking | Insurance, BMI, race/ethnicity | Race/ethnicity, education |
| Diet | Race/ethnicity, BMI | Race/ethnicity, education |
| Sleep | Insurance, BMI, race/ethnicity | Education |
| ***sEng*** |  |  |
| Composite CVH Score | Age, income | Age, education, race/ethnicity, discrimination |
| BMI | Discrimination | Age, education, race/ethnicity, discrimination |
| Blood Pressure | Discrimination, BMI, resilience | Age, BMI, education, race/ethnicity, discrimination |
| Physical Activity | Discrimination, BMI, resilience, race/ethnicity | Age, BMI, education, race/ethnicity, discrimination |
| Smoking | Discrimination, BMI | Age, BMI, education, race/ethnicity, discrimination |
| Diet | Race/ethnicity, BMI | Age, BMI, education, race/ethnicity, discrimination |
| Sleep | Discrimination, BMI, race/ethnicity | Age, BMI, education, race/ethnicity, discrimination |

*BMI: body mass index; PAPP-A: pregnancy-associated plasma protein A; PlGF: placental growth factor; sEng: soluble endoglin; sFlt-1: soluble FMS-like tyrosine kinase 1; VEGF: vascular endothelial growth factor.*

**Table S3.** Descriptive characteristics of full versus selected sample. Data presented as mean ± SD or frequency (%). Means, SD, and percentages for the selected sample are weighted by sampling proportion. Hypothesis tests were conducted using t-tests or Chi-square tests, as appropriate. Significance accepted at p<0.05.

|  | Full sample (N=10,038) | Selected sample (n=2,188) | p-value |
| --- | --- | --- | --- |
| **Maternal age (years)** | 26.9 ± 5.67 | 26.9 ± 5.65 | 0.88 |
| **BMI (kg/m<sup>2</sup>)** | 26.3 ± 6.33 | 26.0 ± 6.10 | **0.02** |
| **Maternal race** |  |  | **0.01** |
| Asian | 407 (4%) | 73 (3%) |  |
| Black non-Hispanic | 1418 (14%) | 352 (13%) |  |
| Hispanic | 1700 (17%) | 360 (16%) |  |
| White non-Hispanic | 5989 (60%) | 1277 (63%) |  |
| Other | 514 (5%) | 125 (5%) |  |
| **Educational Attainment** |  |  | 0.81 |
| Less than high school | 816 (8%) | 218 (8%) |  |
| Completed high school or GED | 1171 (12%) | 289 (12%) |  |
| Some college | 1948 (19%) | 446 (19%) |  |
| Associate or technical degree | 1005 (10%) | 233 (10%) |  |
| Completed college | 2772 (28%) | 541 (27%) |  |
| Degree work beyond college | 2308 (23%) | 460 (24%) |  |
| **Percentage of Federal Poverty Level** |  |  | 0.17 |
| <100% | 1297 (16%) | 298 (15%) |  |
| 100-200% | 1169 (14%) | 255 (13%) |  |
| >200% | 5662 (70%) | 1164 (72%) |  |
| **Insurance payor** |  |  | **0.001** |
| Commercial | 5239 (53%) | 1128 (56%) |  |
| Medicare/Medicaid | 2711 (27%) | 619 (24%) |  |
| Military/Other | 164 (2%) | 31 (1%) |  |
| Two or more sources | 1576 (16%) | 337 (17%) |  |
| None (personal income) | 269 (3%) | 48 (2%) |  |
| **Perceived racial discrimination** |  |  | **<0.001** |
| None | 7172 (71%) | 1621 (76%) |  |
| Moderate | 1587 (16%) | 397 (17%) |  |
| High | 1279 (13%) | 170 (7%) |  |
| **Resilience** | 79.3 ± 11.74 | 79.7 ± 11.84 | 0.16 |
| **APO** | 1502 (15%) | 1349 (16%) | 0.27 |

*APO: adverse pregnancy outcome; BMI: body mass index; GED: general education diploma.*

**Table S4.** Weighted absolute placental biomarker concentrations in each trimester by cardiovascular health categories. Data presented as weighted median and interquartile range.

|  | High CVH (n=495) | Moderate CVH (n=916) | Poor CVH (n=220) | p-value |
| --- | --- | --- | --- | --- |
| **First trimester biomarker concentrations** |  |  |  |  |
| PAPP-A (mU/mL) | 1156.4<br>(565.7 – 2004.0) | 975.0<br>(466.2 – 1972.4) | 650.9<br>(322.1 – 1598.8) | 0.038 |
| VEGF (pg/mL) | 0.72<br>(0.50 – 1.05) | 0.74<br>(0.52 – 1.15) | 0.85<br>(0.56 – 1.62) | 0.777 |
| PlGF (pg/mL) | 44.5<br>(29.5 – 59.3) | 41.2<br>(29.9 – 58.9) | 43.3<br>(29.4 – 60.0) | 0.193 |
| sFlt-1 (pg/mL) | 970.5<br>(711.2 – 1215.4) | 901.9<br>(704.4 – 1182.9) | 841.8<br>(643.8 – 1071.1) | 0.028 |
| sEng (ng/mL) | 6.77<br>(5.77 – 7.83) | 6.27<br>(5.42 – 7.33) | 5.43<br>(4.77 – 6.69) | **<0.001** |
| **Second trimester biomarker concentrations** |  |  |  |  |
| PAPP-A (mU/mL) | 9662.3<br>(5645.7 – 15111.4) | 8867.7<br>(5369.1 – 14644.7) | 7487.5<br>(3436.1 – 11521.9) | **<0.001** |
| VEGF (pg/mL) | 0.92<br>(0.63 – 1.53) | 0.95<br>(0.66 – 1.51) | 1.08<br>(0.72 – 1.97) | 0.112 |
| PlGF (pg/mL) | 203.1<br>(139.8 – 283.5) | 185.4<br>(132.2 – 282.1) | 187.9<br>(121.9 – 302.6) | 0.960 |
| sFlt-1 (pg/mL) | 866.9<br>(624.9 – 1169.7) | 865.5<br>(645.6 – 1215.7) | 857.1<br>(648.4 – 1147.2) | 0.671 |
| sEng (ng/mL) | 5.57<br>(4.89 – 6.36) | 5.29<br>(4.62 – 6.16) | 4.96<br>(4.02 – 5.75) | **<0.001** |

*PAPP-A: pregnancy-associated plasma protein A; PlGF: placental growth factor; sEng: soluble endoglin; sFlt-1: soluble FMS-like tyrosine kinase 1; VEGF: vascular endothelial growth factor*

**Table S5.** Adjusted marginal means (95% confidence intervals) between physical activity categories and 1^st^ and 2^nd^ trimester placental analytes.

|  | Adjusted Marginal<br>Mean (95% CI)<br>Model 1 | p-value | Adjusted Marginal<br>Mean (95% CI)<br>Model 2 | p-value |
| --- | --- | --- | --- | --- |
| **First trimester biomarkers** |  |  |  |  |
| **Log PAPP-A (mU/mL)** |  |  |  |  |
| None | 2.91 (2.87, 2.96) | Reference | 2.92 (2.86, 2.97) | Reference |
| Insufficient | 2.92 (2.88, 2.96) | 0.822 | 2.93 (2.88, 2.97) | 0.813 |
| Sufficient | 2.96 (2.91, 3.01) | 0.177 | 2.93 (2.88, 2.99) | 0.727 |
| High | 2.84 (2.77, 2.92) | 0.130 | 2.81 (2.72, 2.89) | 0.040 |
| **Log VEGF (pg/mL)** |  |  |  |  |
| None | -0.14 (-0.18, -0.10) | Reference | -0.19 (-0.23, -0.14) | Reference |
| Insufficient | -0.09 (-0.14, -0.05) | 0.115 | **-0.08 (-0.13, -0.03)** | **0.002** |
| Sufficient | -0.13 (-0.18, -0.08) | 0.731 | -0.11 (-0.16, -0.06) | 0.039 |
| High | -0.06 (-0.12, 0.002) | 0.041 | **-0.06 (-0.12, 0.01)** | **0.002** |
| **Log PIGF (pg/mL)** |  |  |  |  |
| None | 1.60 (1.58, 1.63) | Reference | 1.58 (1.54, 1.61) | Reference |
| Insufficient | 1.61 (1.59, 1.64) | 0.580 | 1.61 (1.58, 1.64) | 0.157 |
| Sufficient | 1.61 (1.58, 1.64) | 0.649 | 1.61 (1.57, 1.65) | 0.220 |
| High | 1.60 (1.57, 1.63) | 0.982 | 1.61 (1.58, 1.64) | 0.219 |
| **Log sFlt-1 (pg/mL)** |  |  |  |  |
| None | 2.96 (2.94, 2.98) | Reference | 2.98 (2.96, 3.00) | Reference |
| Insufficient | 2.97 (2.95, 2.99) | 0.478 | 2.97 (2.95, 2.99) | 0.669 |
| Sufficient | 2.97 (2.95, 3.00) | 0.475 | 2.97 (2.95, 3.00) | 0.706 |
| High | 2.94 (2.91, 2.96) | 0.117 | **2.92 (2.90, 2.95)** | **0.003** |
| **sEng (ng/mL)** |  |  |  |  |
| None | 6.45 (6.28, 6.62) | Reference | 6.60 (6.40, 6.79) | Reference |
| Insufficient | 6.60 (6.43, 6.78) | 0.225 | 6.65 (6.48, 6.82) | 0.655 |
| Sufficient | 6.59 (6.38, 6.79) | 0.322 | 6.48 (6.27, 6.70) | 0.438 |
| High | 6.61 (6.39, 6.83) | 0.251 | 6.58 (6.35, 6.80) | 0.904 |
| **Second trimester biomarkers** |  |  |  |  |
| **Log PAPP-A (mU/mL)** |  |  |  |  |
| None | 3.98 (3.95, 4.00) | Reference | 3.98 (3.95, 4.01) | Reference |
| Insufficient | **3.92 (3.90, 3.95)** | **0.005** | **3.92 (3.89, 3.94)** | **0.003** |
| Sufficient | 3.93 (3.89, 3.96) | 0.022 | **3.92 (3.89, 3.95)** | **0.006** |
| High | 3.94 (3.91, 3.97) | 0.081 | **3.91 (3.88, 3.95)** | **0.008** |
| **Log VEGF (pg/mL)** |  |  |  |  |
| None | 0.02 (-0.01, 0.05) | Reference | 0.009 (-0.029, 0.048) | Reference |
| Insufficient | 0.02 (-0.01, 0.05) | 0.982 | 0.01 (-0.02, 0.05) | 0.879 |
| Sufficient | 0.04 (0.004, 0.081) | 0.403 | 0.044 (-0.001, 0.089) | 0.253 |
| High | 0.02 (-0.02, 0.06) | 0.858 | 0.02 (-0.03, 0.06) | 0.813 |
| **Log PIGF (pg/mL)** |  |  |  |  |
| None | 2.27 (2.23, 2.30) | Reference | 2.28 (2.24, 2.32) | Reference |
| Insufficient | 2.25 (2.22, 2.28) | 0.558 | 2.24 (2.21, 2.71) | 0.159 |
| Sufficient | 2.27 (2.23, 2.31) | 0.856 | 2.25 (2.20, 2.30) | 0.454 |
| High | 2.26 (2.22, 2.30) | 0.783 | 2.23 (2.18, 2.27) | 0.129 |
| **Log sFlt-1 (pg/mL)** |  |  |  |  |
| None | 2.96 (2.95, 2.97) | Reference | 2.97 (2.95, 2.98) | Reference |
| Insufficient | 2.94 (2.92, 2.95) | 0.031 | 2.94 (2.92, 2.95) | 0.011 |
| Sufficient | 2.94 (2.93, 2.96) | 0.092 | 2.95 (2.93, 2.97) | 0.216 |
| High | 2.95 (2.93, 2.97) | 0.533 | 2.95 (2.93, 2.97) | 0.258 |
| **sEng (ng/mL)** |  |  |  |  |
| None | 5.51 (5.42, 5.60) | Reference | 5.57 (5.46, 5.68) | Reference |
| Insufficient | 5.62 (5.52, 5.73) | 0.100 | 5.67 (5.56, 5.79) | 0.215 |
| Sufficient | 5.55 (5.44, 5.66) | 0.585 | 5.59 (5.46, 5.71) | 0.855 |
| High | 5.58 (5.46, 5.70) | 0.351 | 5.57 (5.45, 5.69) | 0.996 |

*Physical activity categories defined as: None = 0 minutes/week; Insufficient = 1-149 minutes/week; Sufficient = 150-299 minutes/week; High = $\geq 300$ minutes per week. Model 1 adjusted for gestational age at blood draw and 1<sup>st</sup> trimester analyte concentration. Model 2 additionally adjusted for maternal age, race, body mass index, educational attainment, income, insurance payor, perceived racial discrimination, and resilience in a backwards stepwise approach. PAPP-A: pregnancy-associated plasma protein A; PlGF: placental growth factor; sEng: soluble endoglin; sFlt-1: soluble FMS-like tyrosine kinase 1; VEGF: vascular endothelial growth factor.*

